# Identification of Familial Hodgkin Lymphoma Predisposing Genes Using Whole Genome Sequencing

**DOI:** 10.1101/2019.12.12.19014324

**Authors:** Aayushi Srivastava, Sara Giangiobbe, Abhishek Kumar, Dagmara Dymerska, Wolfgang Behnisch, Mathias Witzens-Harig, Jan Lubinski, Kari Hemminki, Asta Försti, Obul Reddy Bandapalli

**Author notes:** **Correspondence:** Obul Reddy Bandapalli. These authors contributed equally (Aayushi Srivastava, Sara Giangiobbe).

## Abstract

Hodgkin lymphoma (HL) is a lymphoproliferative malignancy of B-cell origin that accounts for 10% of all lymphomas. Despite evidence suggesting strong familial clustering of HL, there is no clear understanding of the contribution of genes predisposing to HL. In this study, whole genome sequencing (WGS) was performed on 7 affected and 9 unaffected family members from three HL-prone families and variants were prioritized using our Familial Cancer Variant Prioritization Pipeline (FCVPPv2). WGS identified a total of 98564, 170550 and 113654 variants which were reduced by pedigree-based filtering to 18158, 465 and 26465 in families I, II and III, respectively. In addition to variants affecting amino acid sequences, variants in promoters, enhancers, transcription factors binding sites and microRNA seed sequences were identified from upstream, downstream, 5’ and 3’ untranslated regions. A panel of 565 cancer predisposing and other cancer-related genes and of 2383 high-risk HL genes were also screened in these families to aid further prioritization. Pathway analysis of segregating genes with CADD (Combined Annotation Dependent Depletion Tool) scores > 20 was performed using Ingenuity Pathway Analysis software which implicated several candidate genes in pathways involved in B-cell activation and proliferation and in the network of “Cancer, Hematological disease and Immunological Disease”. We used the FCVPPv2 for further *in silico* analysis and prioritized 45 coding and 79 non-coding variants from the three families. Further literature-based analysis allowed us to constrict this list to one rare germline variant each in families I and II and two in family III. Functional studies were conducted on the candidate from family I in a previous study, resulting in the identification and functional validation of a novel heterozygous missense variant in the tumor suppressor gene *DICER1* as potential HL predisposition factor. We aim to identify the individual genes responsible for predisposition in the remaining two families and will functionally validate these in further studies.

## 1 Introduction

Hodgkin lymphoma (HL) is a lymphoproliferative malignancy originated in germinal center B-cells and is reported to account for about 10% of newly diagnosed lymphomas and 1% of all de novo neoplasms worldwide with an incidence of about 3 cases per 100,000 people in Western countries (Diehl et al., 2004). It is the one of the most common tumors in young adults in economically developed countries, with one peak of incidence in the third decade of life and a second peak after 50 years of age.

Based on differences in the morphology and phenotype of the lymphoma cells and the composition of the cellular infiltrate, HL is subdivided into classical Hodgkin lymphoma (cHL) that accounts for about 95% of cases and nodular lymphocyte-predominant Hodgkin lymphoma (NLPHL) that accounts for the remaining 5% of cases (Kuppers, 2009).

Although familial risk for HL is reported to be among the highest of all cancers (Kharazmi et al., 2015), not many genetic risk factors have been identified. An association between various HLA class I and class II alleles and increased risk of HL has been reported (Diepstra et al., 2005), while other non-HLA susceptibility loci have been detected through genome-wide association studies (Frampton et al., 2013;Cozen et al., 2014;Kushekhar et al., 2014).The identification of major predisposing genes is a more daunting task; however, rare germline variants in *KLDHC8B, NPAT, ACAN, KDR, DICER1* and *POT1* gene have been reported by different groups in high-risk HL families (Salipante et al., 2009;Saarinen et al., 2011;Ristolainen et al., 2015;Rotunno et al., 2016;Bandapalli et al., 65 2018;McMaster et al., 2018).

Here we report the results of whole genome sequencing (WGS) performed in three families with documented recurrence of HL. We used our Familial Cancer Variant Prioritization Pipeline (FCVPPv2) (Kumar et al., 2018) as well as two gene/variant panels based on cancer predisposing genes and variants prioritized in the largest familial HL cohort study to date in order to identify possible disease-causing high-penetrance germline variants in each family (Zhang et al., 2015;Rotunno et al., 2016). Pathway and network analyses using Ingenuity Pathway Analysis software also allowed us to gain insight into the molecular mechanisms of the pathogenesis of HL. We hope that these results can be used in the development of targeted therapy and in the screening of other individuals at risk of developing HL.

## 2 Materials and Methods

### 2.1 Patient samples

Three families with documented recurrence of HL were analyzed in this study, with a total number of 16 individuals (7 affected and 9 unaffected). HL family I and family III were recruited at the University Hospital of Heidelberg, Germany, while family II was recruited at the Pomeranian Medical University, Szczecin, Poland.

The study was approved by the Ethics Committee of the University of Heidelberg and Pomeranian Medical University, Poland. Collection of blood samples and clinical information from subjects was undertaken with a written informed consent in accordance with the tenets of the Declaration of Helsinki.

Germline DNA samples used for genome sequencing were isolated from peripheral blood using QIAamp® DNA Mini kit (Qiagen, Cat No. 51104) according to the manufacturer’s instructions.

### 2.2 Whole genome sequencing, variant calling, annotation and filtering

Whole genome sequencing (WGS) of available affected and unaffected members of the three HL families was performed using Illumina-based small read sequencing. Mapping to reference human genome (assembly version Hs37d5) was performed using BWA mem (version 0.7.8) (Li and Durbin, 2009) and duplicates were removed using biobambam (version 0.0.148). The SAMtools suite (Li, 2011) was used to detect single nucleotide variants (SNVs) and Platypus (Rimmer et al., 2014) to detect indels. Variants were annotated using ANNOVAR, 1000 Genomes, dbSNP and ExAC (Smigielski et al., 2000;Wang et al., 2010;The Genomes Project et al., 2015;Lek et al., 2016).

Variants with a quality score greater than 20 and a coverage greater than 5x, SNVs that passed the strand bias filter (a minimum one read support from both forward and reverse strand) and indels that passed all the Platypus internal filters were evaluated further for minor allele frequencies (MAFs) with respect to the 1000 Genomes Phase 3 and non-TCGA ExAC data. Variants with a MAF less than 0.1 % were deduced from these two datasets. A pairwise comparison of shared rare variants was performed to check for sample swaps and family relatedness.

### 2.3 Data analysis and variant prioritization

#### 2.3.1 Prioritization of coding variants

Variant evaluation was performed using the criteria of our in-house developed variant prioritization pipeline (FCVPPv2) (Kumar et al., 2018). First, variants with MAF < 0.1% were filtered based on the pedigree data considering cancer patients as cases and unaffected persons as controls. The probability of an individual being a Mendelian case or true control was considered.

Variants were then ranked using the combined annotation dependent depletion (CADD) tool v1.3 (Kircher et al., 2014). Only variants with a scaled PHRED-like Combined Annotation Dependent Depletion (CADD) score greater than 10, i.e. variants belonging to the top 1 % of probable deleterious variants in the human genome, were considered further. Genomic Evolutionary Rate Profiling (GERP) (Cooper et al., 2005), PhastCons (Siepel et al., 2005) and PhyloP (Pollard et al., 2010) were used to evaluate the evolutionary conservation of a particular variant. GERP scores > 2.0, PhastCons scores > 0.3 and PhyloP scores ≥ 3.0 were indicative of a good level of conservation and were therefore used as thresholds in the selection of potentially causative variants.

Next, all missense variants were assessed for deleteriousness using 10 tools accessed using dbNSFP (Liu et al., 2016), namely SIFT, PolyPhen V2-HDV, PolyPhen V2-HVAR, LRT, MutationTaster, Mutation Assessor, FATHMM, MetaSVM, MetLR and PROVEAN. Variants predicted to be deleterious by at least 60% of these tools were analyzed further. Prediction scores for nonsense variants were attained via VarSome (Kopanos et al., 2018), the final verdict on pathogenicity offered by VarSome was based on the following tools: DANN, MutationTaster, FATHMM-MKL, FATHMM-XF, ALoFT, EIGEN, EIGEN PC and PrimateAI.

Lastly, three different intolerance scores derived from NHLBI-ESP6500 (Petrovski et al., 2013), ExAC (Lek et al., 2016) and a local dataset, all of which were developed with allele frequency data, were included to evaluate the intolerance of genes to functional mutations. However, these scores were used merely to rank the variants and not as cut-offs for selection. The ExAC consortium has developed two additional scoring systems using large-scale exome sequencing data including intolerance scores (pLI) for loss-of-function variants and Z-scores for missense and synonymous variants. These were used for nonsense and missense variants, respectively. 129

#### 2.3.2 Analysis of non-coding variants

Variants located in the 3’ and 5’ untranslated regions (UTRs) were prioritized based on their location in regulatory regions. Putative miRNA targets at variant positions within 3’ UTRs and 1 kb downstream of transcription end sites were detected by scanning the entire dataset of the human miRNA target atlas from TargetScan 7.0 (Agarwal et al., 2015) using the intersect function of bedtools. Similarly, 5’UTRs and regions 1 kb upstream of transcription start sites were scanned for putative enhancers and promoters using merged enhancer and promoter data from the FANTOM5 consortium as well as super-enhancer data from the super-enhancer archive (SEA) and dbSUPER. These regions were also scanned for transcription factor binding sites using SNPnexus (Dayem Ullah 139 et al., 2018).

The regulatory nature and the possible functional effects of non-coding variants were evaluated using CADD v1.3, HaploReg V4 (Ward and Kellis, 2012) and RegulomeDB (Boyle et al., 2012), primarily based on ENCODE data (Birney et al., 2007). Epigenomic data and marks from 127 cell lines from the NIH Roadmap Epigenomics Mapping Consortium were accessed via CADD v1.3, which gave us information on chromatin states from ChromHmm and Segway. CADD also provided mirSVR scores to rank predicted microRNA target sites by a down-regulation score. These scores are based on a new machine learning method based on sequence and contextual features extracted from miRanda-predicted target sites (Betel et al., 2010). Furthermore, SNPnexus was used to access non-coding scores for each variant and to identify regulatory variants located in CpG islands.

The final selection of 3’UTR and downstream variants was based on their CADD scores > 10 and whether or not they had predicted miRNA target site matches. Similarly, upstream and 5’ UTR variants in enhancers, promoters, super-enhancers or transcription factor binding sites with CADD scores > 10 were short-listed.

#### 2.3.3 Presence of candidate variants in 565 cancer predisposing and other cancer-related genes

In a study on cancer predisposing genes (CPGs) in pediatric cancers, Zhang et al. compiled 565 CPGs based on review of the American College of Medical Genetics and Genomics (ACMG) and medical literature (Zhang et al., 2015). The categories included genes associated with autosomal dominant cancer-predisposition syndromes (60), genes associated with autosomal recessive cancer-predisposition syndromes (29), tumor-suppressor genes (58), tyrosine kinase genes (23) and other cancer genes (395). We investigated a list of genes corresponding to our shortlisted coding and non-coding variants for their presence in the list of genes in the aforementioned study.

#### 2.3.4 Presence of candidate variants in prioritized HL genes from a large WES-based familial HL study

In a study by Rotunno et al., 2699 variants corresponding to 2383 genes were identified in 17 HL discovery families after filtering and prioritization (Rotunno et al., 2016). We intersected our list of candidate genes with this list of 2383 HL genes to identify coding and non-coding variants from our shortlist in potentially causative HL genes.

### 2.4 Variant validation

Specific variants of interest mentioned throughout the text were validated using specific primers for polymerase chain reaction amplification designed with Primer3 (http://bioinfo.ut.ee/primer3-0.4.0/) and Sanger sequencing on a 3500 Dx Genetic Analyzer (Life Technologies, CA, USA), using ABI PRISM 3.1 Big Dye terminator chemistry, according to the manufacturer’s instructions. The electrophoretic profiles were analyzed manually. Segregation analysis of the prioritized variants was performed in additional family members when DNA was available. Primer details are available on request.

### 2.5 Ingenuity pathway analysis (IPA)

IPA (Qiagen; http://www.qiagen.com/ingenuity; analysis date 15/10/2019) was used to perform a core analysis to identify enriched canonical pathways, diseases, biological functions and molecular networks among genes that passed the allele frequency cut-off, fulfilled family-based segregation criteria, met the CADD score cut-off and were not intergenic or intronic variants. Data were analyzed for all three families together. Top canonical pathways were identified from the IPA pathway library and ranked according to their significance to our input data. This significance was determined by p-values calculated using the right tailed Fisher’s exact test.

IPA was also used to generate gene networks in which upstream regulators were connected to the input dataset genes while taking advantage of paths that involved more than one link (i.e., through intermediate regulators). These connections represent experimentally observed cause-effect relationships that relate to expression, transcription, activation, molecular modification and transport as well as binding events. The networks were ranked according to scores that were generated by considering the number of focus genes (input data) and the size of the network to approximate the relevance of the network to the original list of focus genes.

## 3 Results

### 3.1 Whole genome sequencing results

In our study, we analyzed three families with reported recurrence of Hodgkin lymphoma. Their respective pedigrees are shown in Figure 1.

**Figure 1.**
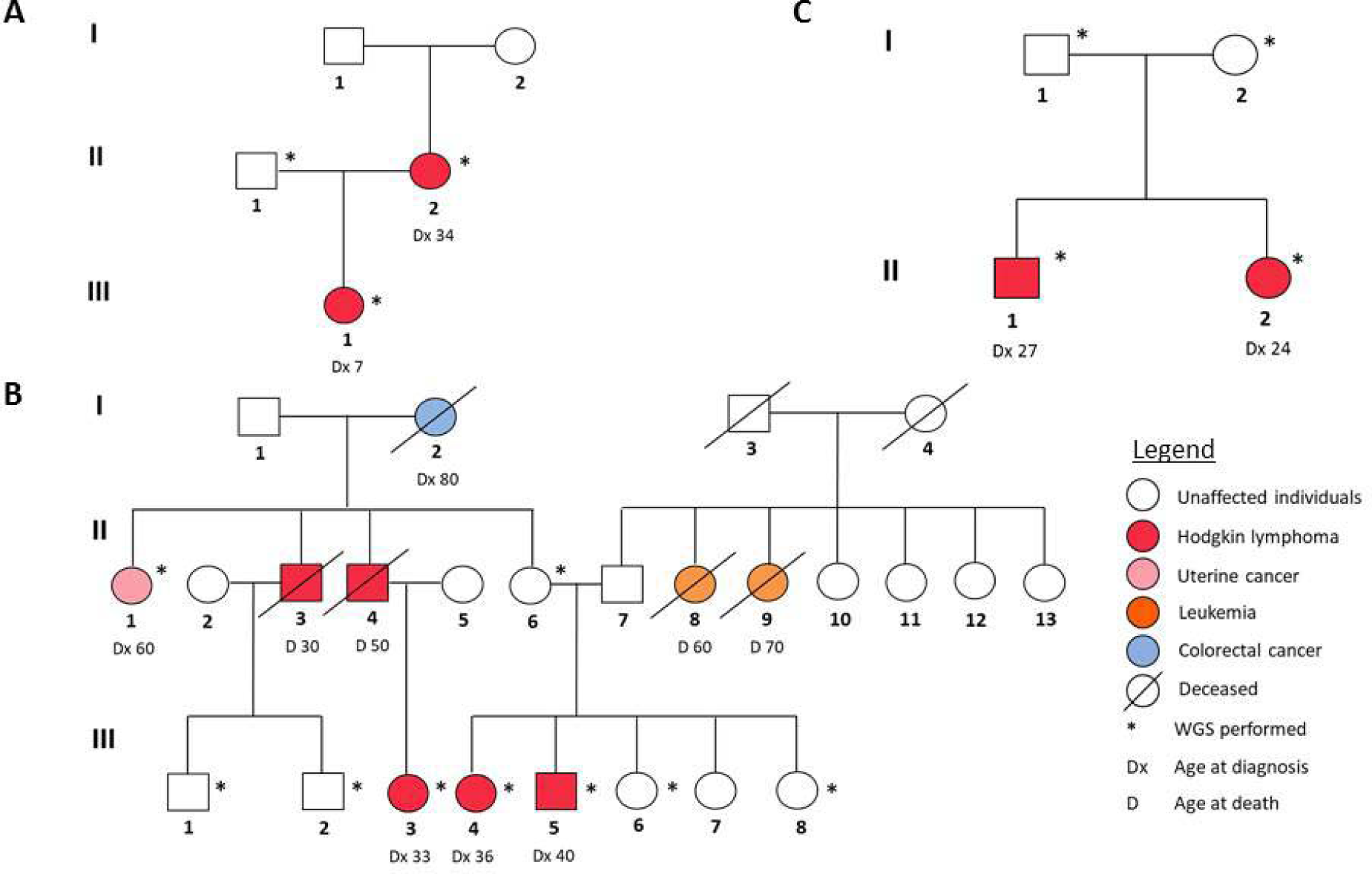
Pedigrees of the three HL families analyzed in this study. A: Family 1, B: Family 2, C: Family 3.

**Figure 2.**
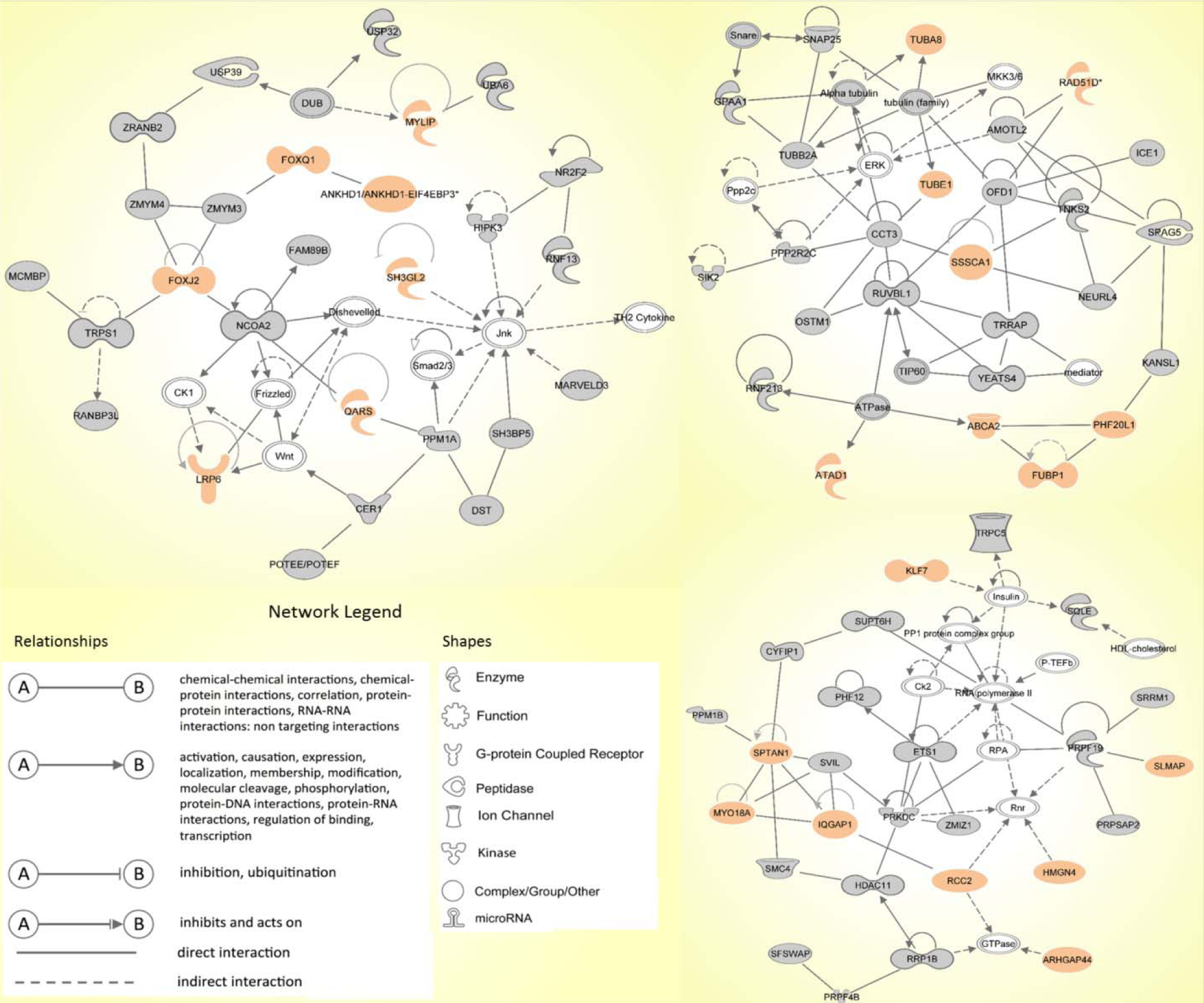
The top three molecular networks identified by Ingenuity Pathway Analysis: (a) Network 1. Cancer, Hematological Disease, Immunological Disease; (b) Network 2. Developmental Disorder, Endocrine System Disorders, Hereditary Disorder; (c) Network 3. RNA Post-Transcriptional Modification, Cell Death and Survival, Cellular Movement. Genes from our input-data are shown in grey, genes from our prioritized candidate list are highlighted in peach.

In family I (Figure 1A), the proband (III-1) and her mother (II-2) were diagnosed with two different histological subtypes of classical Hodgkin lymphoma (cHL) at the ages of 7 and 34, respectively. The daughter was diagnosed with nodular sclerosis cHL and the mother with lymphocyte-rich cHL.

The sample of the unaffected father (II-1) was also sequenced. Family II (Figure 1B) is characterized by a strong recurrence of HL. Five family members were diagnosed with HL (II-3, II-4, III-3, III-4 and III-5), of which three (III-3, III-4 and III-5) underwent WGS. In addition, the family member (II-6), who was considered as an obligatory carrier of the mutation, was sequenced as were samples and four healthy family members (III-1, III-2, III-6 and III-8) and one family member diagnosed with uterine cancer (II-1) as controls. In family 3 (Figure 1C), II-1 and II-2 were diagnosed with cHL, at the age of 27 and 24 respectively. Their parents (I-1, I-2) were not affected, however one of them is expected to be a carrier and analyzed accordingly.

WGS of 7 affected and 9 unaffected members from the three studied families identified a total number of 98564, 170550 and 113654 variants which were reduced by pedigree-based filtering to 18158, 465 and 26465 in families I, II and III, respectively.

### 3.2 Prioritization of candidates according to the FCVPPv2

After pedigree-based filtering, 130, 7 and 196 exonic variants were left in families I, II and III, respectively, with a prevalence of non-synonymous and synonymous SNVs. The predominant type of substitution was the C>T transition. Among exonic variants fulfilling pedigree-based criteria, only variants with CADD scores > 10 were taken into further consideration and prioritized according to deleteriousness, intolerance and conservational scores, as detailed in the methods section. At the end of this process, 37 potential missense variants and 9 potential nonsense mutations were prioritized for families I - III and are shown in Tables 1 and 2.

**Table 1.**
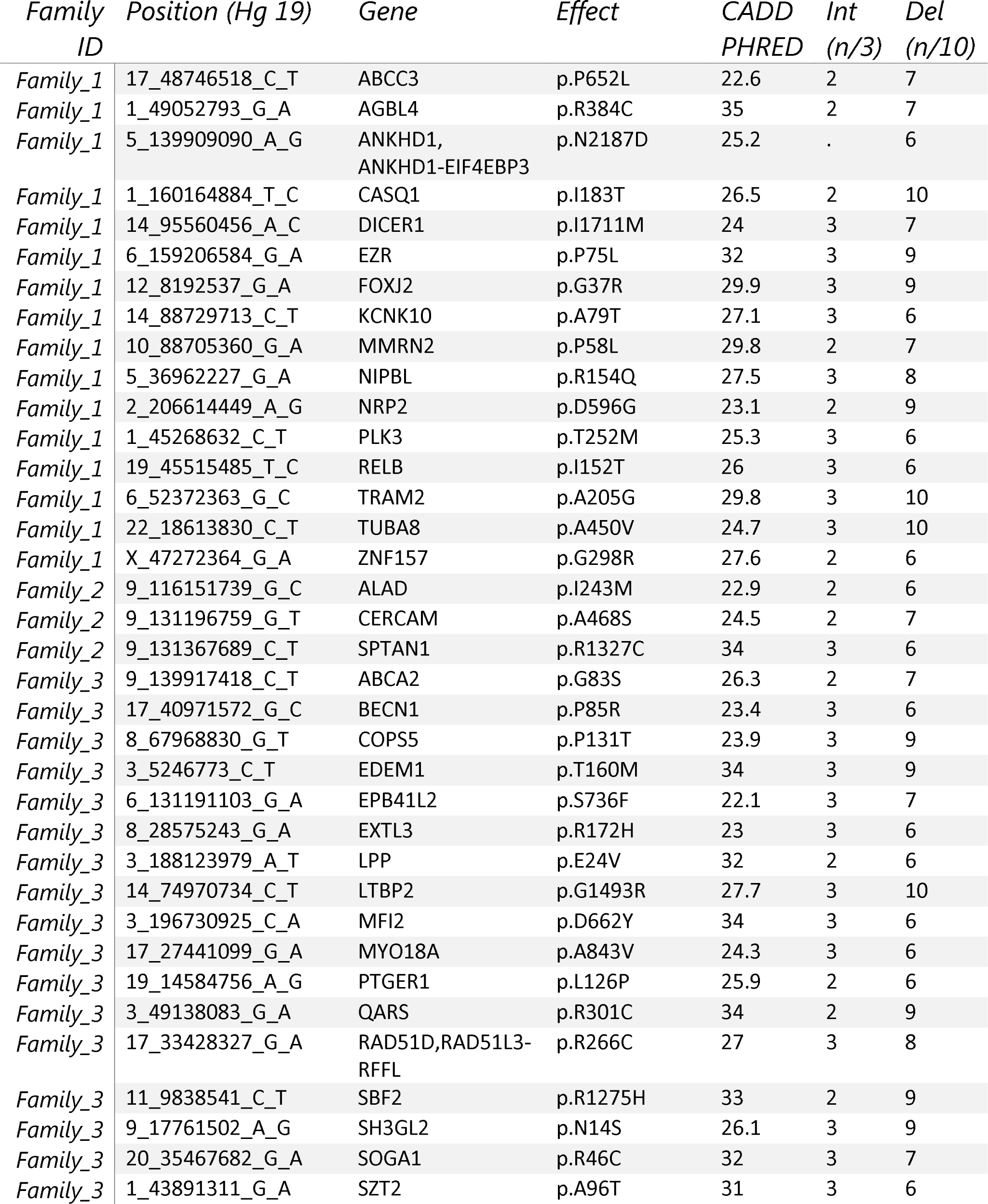
Top <u>missense</u> variants prioritized using the FCVPPv2. Chromosomal positions, classifications, PHRED-like CADD scores, protein changes and the number of positive intolerance (Int) and deleteriousness (Del) scores are shown for each variant.

**Table 2.**
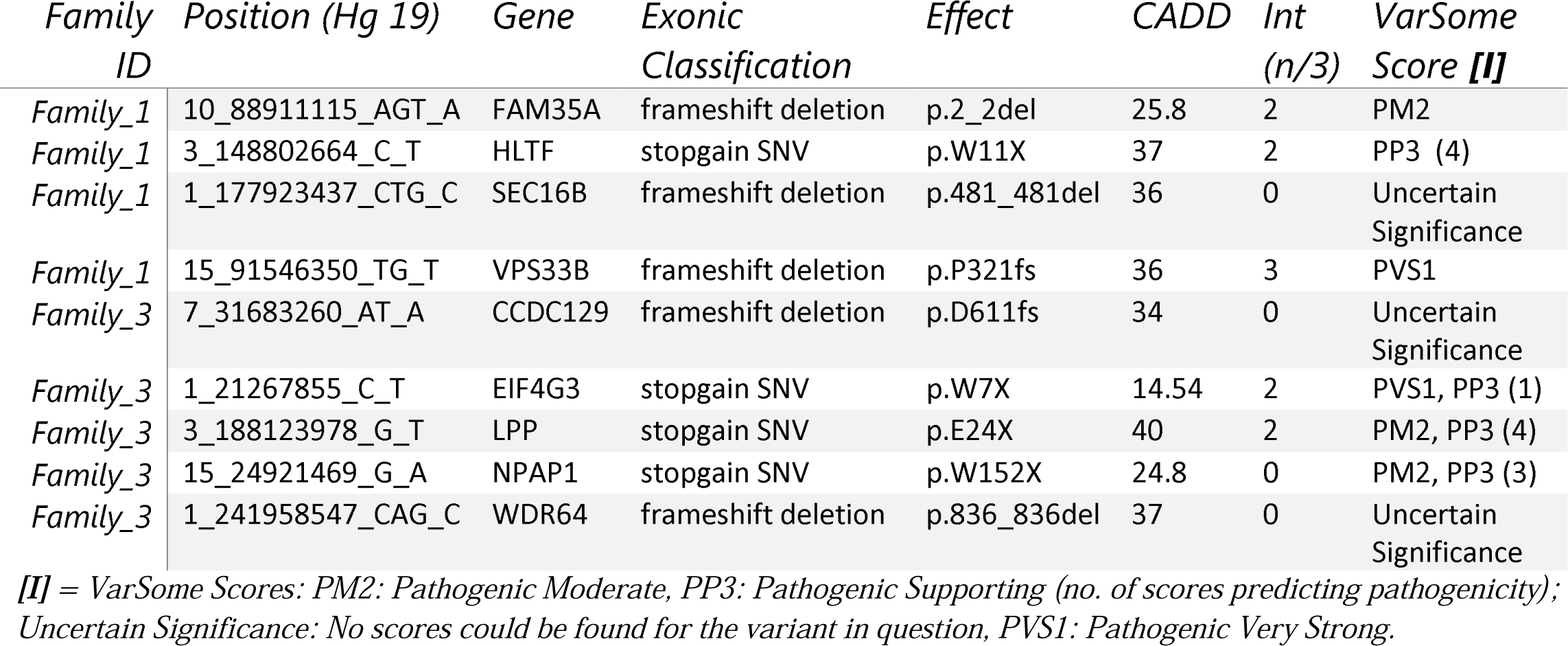
Top <u>non-sense</u> variants prioritized using the FCVPPv2. Chromosomal positions, classifications, PHRED-like CADD scores, protein changes, the number of positive intolerance (Int) and VarSome prediction scores are included for each variant.

Pedigree-based filtering also reduced the number of potentially interesting variants located in the untranslated regions to 523 for 5’UTR variants (130 in family I, 5 in family II and 314 in family III) and 854 for 3’UTR variants (347 in family I, 10 in family II and 497 in family III). These variants were further prioritized based on their CADD score > 10 and their localization in known regulatory regions (Supplementary Table 1). 5’UTR variants were analyzed by the SNPNexus tool, which allowed us to identify 4 variants located in transcription factors binding sites. In addition, the intersect function of bedtools was used to identify further 15 variants located in promoter regions and 4 located in super-enhancer regions. Among variants located in the 3’UTR region, 56 variants located in miRNA seed sequences were selected.

### 3.3 Candidate variants in 565 CPGs and 2383 potentially causative HL genes

Intersecting our prioritized list of candidate genes with the list of 565 CPGs identified 11 genes with variants in coding and selected non-coding regions (upstream and downstream variants, 3’ and 5’ UTRs) of the known CPGs. These include *FUBP1, SEPT6, DICER1, EZR* and *NCOA1* from family 1 and *BCL6, RAD51D, LPP* and *PTCH1* from family 3 (Table 3). *DICER1* and *PTCH1* are known in autosomal dominant cancer-predisposition syndromes, whereas the rest are categorized as being “other cancer genes”.

**Table 3.** Variants corresponding to genes present in the panel of 565 known cancer predisposition genes from a study by Zhang et al.

| Gene ID | HL Family | HL Gene | HL Variant | Variant Type | Variant Classification | HGNC Approved Name | CADD_PHRED | Familial Syndrome | Category |
| --- | --- | --- | --- | --- | --- | --- | --- | --- | --- |
| 10499 | 1 | NCOA2 | 8_71316112_T_TCCTCCTCCC | Indel | upstream | nuclear receptor coactivator 2 | 15.56 |  | Other CancerGene |
| 8880 | 1 | FUBP1 | 1_78414225_A_G | SNVs | UTR3 | far upstream element (FUSE) binding protein 1 | 13.59 |  | Other CancerGene |
| 23157 | 1 | SEPT6 | X_118751062_CGTGT_C | Indel | UTR3 | septin 6 | 10.56 |  | Other CancerGene |
| 23405 | 1 | DICER1 | 14_95560456_A_C | SNVs | nonsynonymous SNV | dicer 1, ribonuclease type III | 24 | DICER1 syndrome, Familial Multinodular Goiter | Autosomal Dominant |
| 7430 | 1 | EZR | 6_159206584_G_A | SNVs | nonsynonymous SNV | ezrin | 32 |  | Other CancerGene |
| 604 | 3 | BCL6 | 3_187463568_C_A | SNVs | upstream; downstream | B-cell CLL/lymphoma 6 | 13 |  | Other CancerGene |
| 4026 | 3 | LPP | 3_188123978_G_T | SNVs | stopgain SNV | LIM domain containing preferred translocation partner in lipoma | 40 |  | Other CancerGene |
|  | 3 | LPP | 3_188123979_A_T | SNVs | nonsynonymous SNV | LIM domain containing preferred translocation partner in lipoma | 32 |  | Other CancerGene |
|  | 3 | LPP | 3_188608373_A_T | SNVs | UTR3 | LIM domain containing preferred translocation partner in lipoma | 10.5 |  | Other CancerGene |
| 5892 | 3 | RAD51D | 17_33428327_G_A | SNVs | nonsynonymous SNV | RAD51 paralog D | 27 |  | Other CancerGene |
| 5727 | 3 | PTCH1 | 9_98270531_C_A | SNVs | nonsynonymous SNV | patched 1 | 20.4 | Gorlin syndrome | Autosomal Dominant |

In addition to the identification of 11 variants in CPGs, we intersected our prioritized list of genes with a list of 2383 genes with potentially causative variants from a large WES-based familial HL study. We found 25 variants in the coding and non-coding regions in 23 of the HL genes, with 7 coming from family I and 18 from family III (Table 4).

**Table 4.**
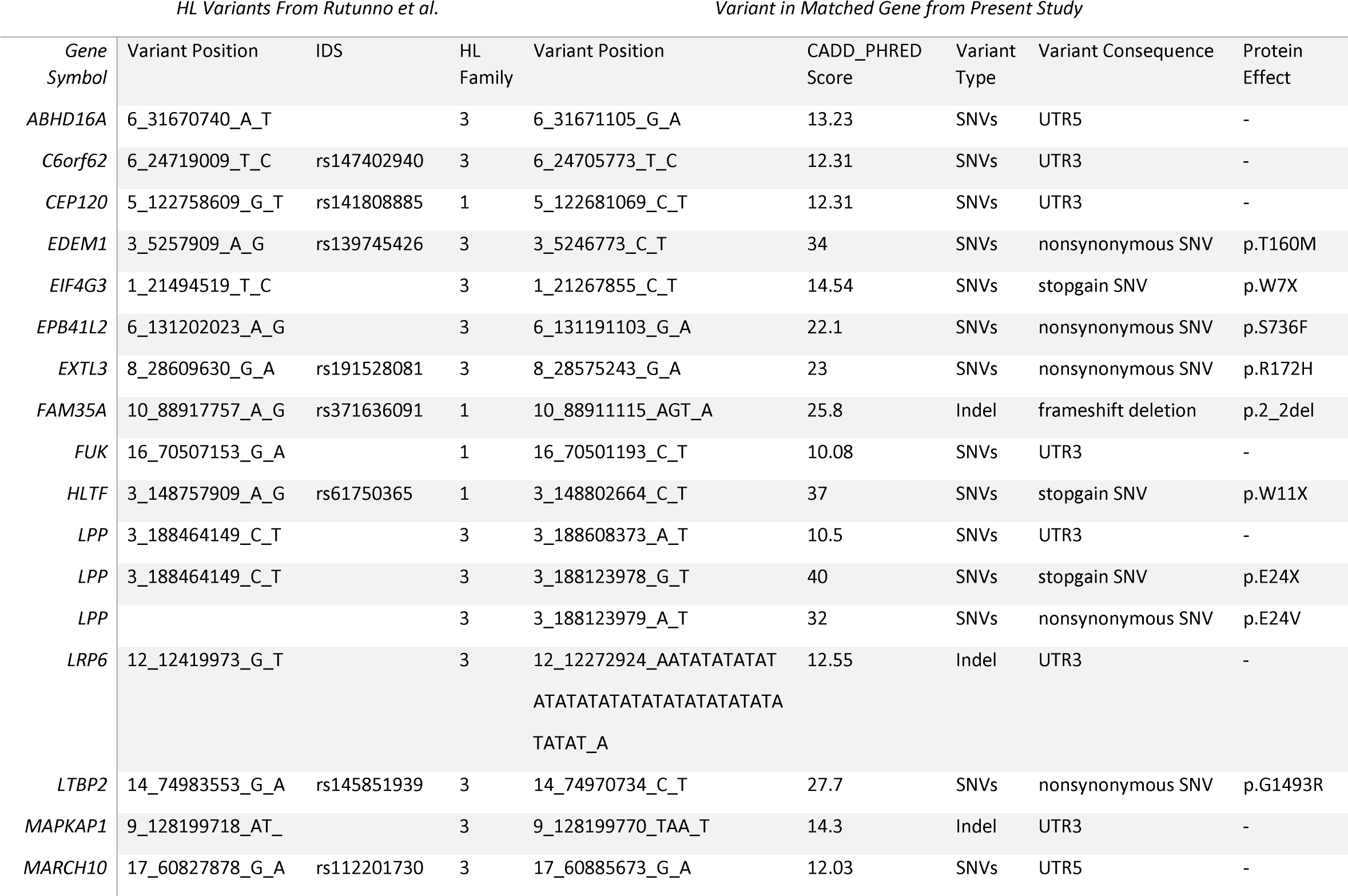
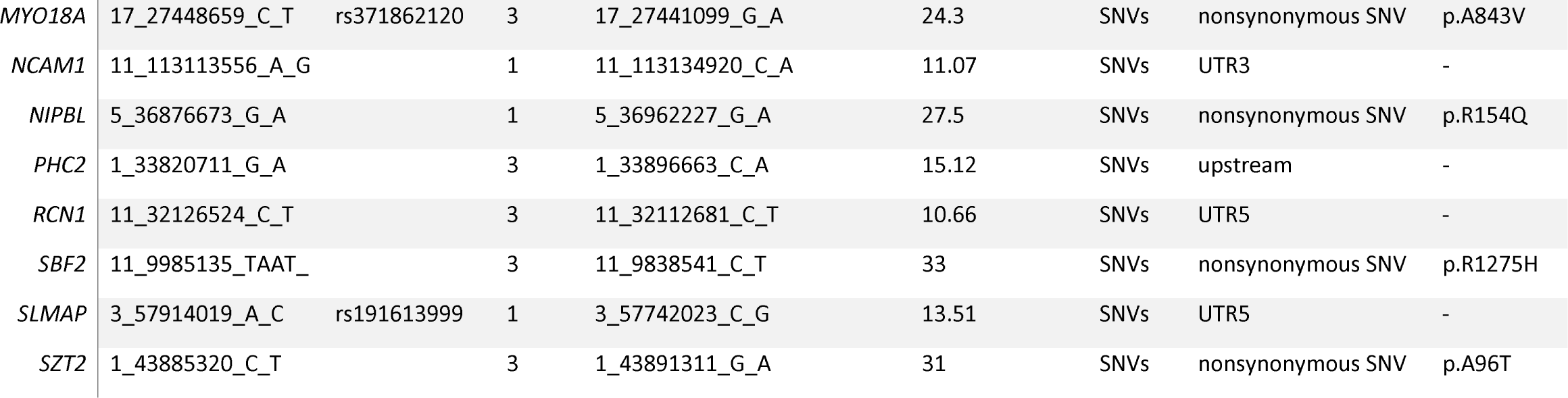
Variants corresponding to genes intersecting with the list of 2383 high-risk HL genes from a study by Rutunno et al. Variant details from both databases (the present study and the study by Rutunno et al.) are shown.

### 3.4 Network and pathway analysis with IPA

Pathway analysis of the selected variants performed with IPA showed an enrichment of mutations in genes involved in pathways essential for B-cell proliferation and activation, specifically B-cell receptor signaling, and PI3K signaling in B lymphocytes and B cell activating factor signaling (Supplementary Table 2A, Supplementary Figures 1A and 1B).

Similarly, the IPA network analysis generated a comprehensive picture of possible gene interactions between our candidate genes (Supplementary Table 2B). The top network is related to cancer, hematological disease and immunological disease, which is in complete coherence with the pathogenesis of HL. Many genes from the prioritized list of top candidates are shown to play a role in the top networks (Figure 2).

### 3.5 Literature mining, consolidation of results and selection of candidates

With the aim of identifying high penetrance dominant variant per family, we used our pipeline results and literature-based mining to determine the genes’ link to Hodgkin lymphoma or immune-related processes. For family 1, we have short-listed 5 potential candidates (*DICER1, HLTF, NOTCH3, PLK3 and RELB*). Based on segregation, confirmation and functional validation, we identified *DICER1* as a candidate HL predisposing gene by showing significant down-regulation of tumor suppressor miRNAs in *DICER1*-mutated family members (Bandapalli et al., 2018). The presence of *DICER1* in the list of 565 CPGs also reinforces its status as the disease-causing variant in this family.

In family 2, three exonic variants made it to the final list (*ALAD, CERCAM* and *SPTAN1*) of which *SPTAN1* was shown to be among the genes in one of the top IPA networks (Network 3; Figure 2C). No coding or non-coding variants intersected with the panel of CPGs or HL candidate genes.

Two genes stand out in family 3, namely *LPP* and *RAD51D*. Both genes were found in the list of 565 CPGs and *LPP* was additionally found in the gene list from the large cohort of HL families. Three variants in *LPP* were prioritized by the FCVPPv2 and made it to the shortlist including one stopgain variant (3_188123978_G_T), one 3’UTR (3_188608373_A_T) and one nonsynonymous missense variant (3_188123979_A_T). LPP (LIM domain containing preferred translocation partner in lipoma) is a member of the zyxin family of LIM proteins that is characterized as a promoter of mesenchymal/fibroblast cell migration. LPP has been shown to be a critical inducer of tumor cell migration, invasion and metastasis by virtue of its ability to localize to adhesions and to promote invadopodia formation (Ngan et al., 2018). A genome-wide association study of 253 Chinese individuals with B-cell NHL also identified a new susceptibility locus between *BCL6* and *LPP* that was significantly associated with the increased risk of B-cell NHL (Tan et al., 2013). On the other hand, there are no reports of an association between *RAD51D* and lymphomas; however, it is a well-established susceptibility gene in Breast-Ovarian Cancer, Familial 4 and Hereditary Breast Ovarian Cancer Syndrome (Loveday et al., 2011;Chen et al., 2018). The final selection of a candidate in this family will be based on further functional studies.

## 4 Discussion

In summary, WGS data analysis of three families with reported recurrence of HL allowed us to prioritize 45 coding and 79 non-coding variants from which we subsequently selected and validated one for family I, (*DICER1*), short-listed three and two for families II (*ALAD, CERCAM* and *SPTAN1*) and III (*RAD51*and *LPP*) respectively to investigate further with validation and functional studies.

For family I we have already functionally validated *DICER1* as the candidate predisposing gene in a previous study (Bandapalli et al., 2018). However, the family was included in this paper, especially with regard to the pathway and network analyses. We identified pathways related to B-cell proliferation and networks related to cancer, hematological disease, immunological disease, hereditary disorders, cell death and cell survival using IPA software, helping us to prioritize genes with functions in the pathogenesis of HL.

Personalized medicine is an upcoming and promising field of medicine in which medical decisions, practices, interventions and products are tailored to the individual patient based on their predicted response or risk of disease. The scope of this field has advanced rapidly with the advent of genomics and other omics and the possibility of implicating one gene or a set of genes in the pathogenesis of a particular disease. Thus, the identification of germline predisposing genes could be of great value in the screening of individuals at risk of developing HL, as well as in the development of personalized adjuvant therapies based on the affected pathways. In this aspect, delta-aminolevulinate dehydratase (*ALAD*) from family 2 is interesting as it is involved in the catalysis of second step in the biosynthesis of heme and is also acts as an endogenous inhibitor of the 26S proteasome, a multi-catalytic ATP-dependent protease complex that functions as the degrading arm of the ubiquitin system:the major pathway for regulated degradation of proteins in all eukaryotes. Down regulation of *ALAD* is shown to be associated with poor prognosis in patients with breast cancer (Ge et al., 2017) whereas the existing data on nonerythroid spectrin αII (*SPTAN1*) suggest that overexpression of *SPTAN1* in tumor cells reflects neoplastic and tumor promoting activity or tumor suppressing effects by enabling DNA repair through interaction with DNA repair proteins (Ackermann and Brieger, 2019) and *CERCAM* is known as an unfavourable prognostic marker in urothelial, rencal and ovarian cancers implying the importance of the variants in these genes (Ma et al., 2016). *RAD51D* from family III is particularly interesting since RAD51D is involved in DNA repair through homologous recombination. Therefore, it is possible that carcinomas arising in patients carrying mutations in this gene will be sensitive to chemotherapeutic agents that target this pathway, such as cisplatin and the PARP (poly (ADP-ribose) polymerase) inhibitor olaparib. This has already been demonstrated in *BRCA1/2* mutation-carrier cancer patients (Banerjee et al., 2010;Loveday et al., 2011). This approach can also be applied to target pathways affected by the mutated genes. Several candidate genes were identified by IPA pathway analysis in B cell receptor pathways, offering a valuable target for other pharmaceutical drugs. The B cell receptor (BCR) signaling pathway, when dysregulated, is a potent contributor to lympomagenesis and tumor survival (Valla et al., 2018). This pathway has been targeted in B-cell lymphomas and leukemias with several BCR-directed agents, such as inhibitors of Bruton’s tyrosine kinase (BTK9), spleen tyrosine kinase (SYK) and phosphatidylinositol-3-kinase (PI3K) (Buggy and Elias, 2012;Dreyling et al., 2017;Liu and Mamorska-Dyga, 2017). In one study, excellent response rates could be demonstrated in certain non-Hodgkin lymphoma subtypes, however, issues related to the development of resistance to BTK inhibitors need to be addressed (Valla et al., 2018).

Advancements in the field of genomics have allowed WGS to become the state-of-the-art tool for the identification of novel cancer predisposing genes in Mendelian diseases. It is still a challenge to appropriately interpret the immense amount of data generated by WGS, especially with respect to non-coding variants. In our study, we have attempted to interpret a selection of non-coding variants using *in-silico* and bioinformatic tools, however, the adequate analysis of intronic and intergenic variants remains a challenge. There are several reports of WGS being successfully implemented to implicate rare, high-penetrance germline variants in cancer, for example *POT1* mutations in familial melanoma and Hodgkin lymphoma (McMaster et al., 2018;Wong et al., 2019) and *POLE* and *POLD1* mutations in colorectal adenomas or carcinomas (Palles et al., 2013). In a previous study, we have used our pipeline (FCVPPv2) to prioritize novel variants in non-medullary thyroid cancer prone families (Srivastava et al., 2019). We have also successfully combined our pipeline with literature review and functional studies to identify *DICER1* as a candidate predisposing gene in one Hodgkin lymphoma family (Bandapalli et al., 2018). We aim to apply these methods in the remaining Hodgkin lymphoma families and hope that these results will facilitate personalized therapy in the studied families and contribute to the screening of other individuals at risk of developing HL.

## Data Availability

Unfortunately, we are not able to provide the sequencing data into a public data base. The data underlying the results presented in the study are available from the corresponding author or from Dr. Asta Försti.

## 5 Conflict of Interest

*The authors declare that the research was conducted in the absence of any commercial or financial relationships that could be construed as a potential conflict of interest*.

## 6 Author Contributions

O.R.B., A.F. and K.H. conceived and designed the study. W.B., M.W-H., D.D. and J.L. provided the HL family samples. A.S, O.R.B., S.G. and A.K. analyzed the data. O.R.B. and S.G. performed the experiments. A.S. and O.R.B. wrote the first draft of the manuscript. All authors read, commented on and approved the manuscript.

## 7 Funding

The study was supported by the Harald Huppert Foundation and Transcan ERA-NET funding from the German Federal Ministry of Education and Research (BMBF).

## 8 Acknowledgments

The authors thank the Genomics and Proteomics Core Facility (GPCF) of the German Cancer Research Center (DKFZ) for providing excellent library preparation and sequencing services, the Omics IT and Data Management Core Facility (ODCF) of the DKFZ for the whole genome sequencing data management and Nagarajan Paramasivam and Matthias Schlesner for their help with analysis of the sequencing data.

## 9 Supplementary Material

The Supplementary Material for this article can be found online at:

## Notes

### Competing Interest Statement

The authors have declared no competing interest.

